# Predictive utility of mortality by aging measures at different hierarchical levels and the response to modifiable lifestyle factors: Implications for geroprotective programs

**DOI:** 10.1101/2021.11.27.21266932

**Authors:** Jingyun Zhang, Xingqi Cao, Chen Chen, Liu He, Ziyang Ren, Junhua Xiao, Liyuan Han, Xifeng Wu, Zuyun Liu

## Abstract

**Background:** Aging, as a multi-dimensional process, can be measured at different hierarchical levels including biological, phenotypic, and functional levels. The aims of this study were to: 1) compare the predictive utility of mortality by three aging measures at three hierarchical levels; 2) develop a composite aging measure that integrated aging measures at different hierarchical levels; and 3) evaluate the response of these aging measures to modifiable lifestyle factors.

**Methods:** Data from National Health and Nutrition Examination Survey 1999-2002 were used. Three aging measures included telomere length (TL, biological level), Phenotypic Age (PA, phenotypic level), and frailty index (FI, functional level). Mortality information was collected until Dec. 2015. Cox proportional hazards regression and multiple linear regression models were performed.

**Results:** A total of 3249 participants (20-84 years) were included. Both accelerations (accounting for chronological age) of PA and FI were significantly associated with mortality, with HRs of 1.67 (95% confidence interval [CI] = 1.41-1.98) and 1.59 (95% CI = 1.35-1.87), respectively, while that of TL showed nonsignificant associations. We thus developed a new composite aging measure (named PC1) integrating the accelerations of PA and FI, and demonstrated its better predictive utility relative to each single aging measure. PC1, as well as the accelerations of PA and FI, were responsive to several lifestyle factors.

**Conclusion:** The findings, for the first time, provide a full picture of the predictive utility of mortality by three aging measures at three hierarchical levels and the response to modifiable lifestyle factors, with important implications for geroprotective programs.

## Introduction

Aging is a critical risk factor for many chronic diseases. As a comprehensive and multi-dimensional process, aging could be measured at different hierarchical levels, including biological, phenotypic and functional levels [1]. Biological aging measures focus on changes at the molecular, cellar, and intracellular levels, such as telomere length (TL) and DNA methylation clocks [1-3]. Phenotypic aging measures include composite indexes derived from multi-system clinical chemistry biomarkers, such as Phenotypic Age (PA) [4], reflecting changes in body composition, homeostatic mechanisms, energetics, and brain health over time. Functional aging measures include composite indexes derived from different functional aspects (e.g., cognitive and physical function). Frailty index (FI) is a widely used functional aging measure that integrates deficits across multiple functional domains [5-7]. These aging measures are conceptually different; however, direct comparative analyses of their predictive utility of mortality risk are limited. To the best of our knowledge, only one study based on adults >50 years in Sweden compared aging measures at three hierarchical levels [8]. Since aging starts early in life [9], it remains unclear how these aging measures behaves in terms of mortality prediction among a general population with younger, middle-aged, and older adults. It is also of interest to examine whether integrating two or more aging measures at different hierarchical levels would provide a more informative one, which is valuable in geroprotective programs where these aging measures serve as endpoints to help with assessing the effectiveness of interventions.

One important feature of qualifying aging measures includes effective responsiveness to interventions [10]. This feature has been rarely emphasized in previous work whereas it is the key to the application of aging measures in clinical settings. Lifestyles such as smoking and physical activity are modifiable factors and have been demonstrated to be associated with individual aging measures such as TL [11] and FI [12, 13]. However, few studies have simultaneously evaluated the response of aging measures at different hierarchical levels to modifiable lifestyle factors in the same population.

Using data from the National Health and Nutrition Examination Survey (NHANES) 1999-2002, including three aging measures at three hierarchical levels (i.e., TL, PA, and FI), this study aimed to 1) compare the predictive utility of mortality risk by three aging measures at three hierarchical levels; 2) develop a new composite aging measure that integrated aging measures at different hierarchical levels; and 3) evaluate the response of these aging measures to modifiable lifestyle factors.

## Materials and Methods

### Study population

NHANES is an ongoing program conducted by the National Center for Health Statistics of the Centers for Disease Control and Prevention in the United States. NHANES began in the early 1960s and focuses on the health and nutritional status of adults and children in the United States. Since 1999, NHANES has become a continuous program that collects a wide range of health-related data via interview, examination, and laboratory tests in counties across the country biennially [4]. NHANES is approved by the National Center for Health Statistics Research Ethics Review Board, and all participants provided informed consent. In this study, we included participants with TL data and complete information to calculate PA and FI. In total, 3249 of 9882 participants aged from 20 to 84 years in NHANES 1999-2002 were included. NHANES data are publicly available (https://www.cdc.gov/nchs/nhanes/index.htm). The analytic roadmap of this study is shown in **Fig. S1** (**Supplementary file**).

### Measurements

#### TL and acceleration of TL (TL.Accel)

In NHANES, TL assay was performed in the laboratory of Dr. Elizabeth Blackburn at the University of California, San Francisco, using the quantitative polymerase chain reaction (PCR) method to measure TL relative to standard reference DNA (T/S ratio) based on blood samples [14, 15]. Each sample was assayed three times on three different days. The mean of the T/S ratio was used to represent TL and details of laboratory methods are described at the official website of NHANES (https://www.n.cdc.gov/Nchs/Nhanes/1999-2000/TELO_A.htm). To eliminate the effect of chronological age (CA) on TL, we calculated a new index, TL.Accel, defined as residual from a linear model when regressing TL on CA. TL.Accel was classified into normal (TL.Accel ≥ 0, indicating that a participant’s TL is equal or longer than expected based on his/her CA) or shorter (TL.Accel < 0, indicating that a participant’s TL is shorter than expected based on his/her CA).

#### PA and acceleration of PA (PA.Accel)

PA was first developed based on NHANES III [4, 16]. In brief, PA was derived from CA and 9 biomarkers including albumin, creatinine, glucose, (log) C-reactive protein, lymphocyte percent, mean cell volume, red cell distribution width, alkaline phosphatase, and white blood cell count. As done to TL, we calculated PA.Accel, defined as residual from a linear model when regressing PA on CA. PA.Accel represents phenotypic aging after accounting for CA, i.e., a participant is phenotypically older (younger) if his/her PA.Accel > 0 (< 0) than expected based on his/her CA [4, 16].

#### FI and acceleration of FI (FI.Accel)

FI integrates 36-item deficits (**Table S1** in **Supplementary file**) including comorbidities, activities of daily living, physical tasks, cognition, and performance testing [17]. FI was calculated as a ratio of the number of deficits in a participant out of the total possible deficits considered, with a range of 0 to 1, and the higher score indicates the frailer a participant was. FI.Accel was defined as residual from a linear model when regressing FI on CA. FI.Accel was used as a categorical variable, and divided into frail (FI.Accel > 0) or robust (FI.Accel ≤ 0).

#### Mortality

Mortality follow-up was based on linked data from records taken from the National Death Index (NDI) through December 31, 2015, provided by the Centers for Disease Control and Prevention [18]. Survival time was calculated as months from the date of interview to the date of death or the end of follow-up, whichever came first.

#### Covariates

Demographic factors (CA, gender, ethnicity, and education level), body mass index (BMI), and lifestyle factors (i.e., smoking status, binge drinking status, alcohol consumption, leisure-time physical activity level [PAQ], and health eating index-2010 [HEI-2010] [19]) were included as covariates. Ethnicity was grouped as non-Hispanic white, non-Hispanic black, Hispanic, and others. Education level was grouped as less than high school (< HS), HS/general educational development (HS/GED), having attended college but not receiving at least a bachelor’s degree (some college), and having a bachelor’s degree or higher (college). BMI was grouped as underweight (BMI < 18.5 kg/m^2^), normal (18.5 kg/m^2^ ≤ BMI < 25 kg/m^2^), overweight (25 kg/m^2^ ≤ BMI < 30 kg/m^2^) and obese (BMI ≥ 30 kg/m^2^). Smoking status was grouped as never smoker, former smoker, and current smoker. Alcohol consumption was grouped as never drinker (never drinking or didn’t drink in the past year), low to moderate drinker (drinks less than three times per month), and heavy drinker (drinks at least one time per week). PAQ was grouped as low (< one time per week), moderate (1-2 times per week), and heavy (≥ 3 times per week). HEI-2010 was grouped by tertiles (Tertile 1, 2, and 3).

### Statistical analyses

The basic characteristics are presented as mean ± standard deviation (SD) and number (percentage) for continuous and categorical variables, respectively.

To assess the predictive utilities for all-cause mortality of three aging measures, survival analysis was conducted. Kaplan-Meier (K-M) curves were plotted and log-rank tests were conducted. Meanwhile, Cox proportional hazards regression was performed based on three models: model 1 was a crude model; model 2 adjusted for CA and gender; and model 3 additionally adjusted for ethnicity, education level, smoking status, alcohol consumption, binge drinking status, BMI, PAQ, and HEI-2010 based on model 2. Hazard ratio (HR) and 95% confidence intervals (95% CI) were documented. Next, time-dependent receiver operating characteristic (ROC) curves [20] were applied to evaluate the predictive utility of different aging measures using model 2 and model 3. Three indices of predictive utility (i.e., area under the curve [AUC], integrated discrimination improvement [IDI], and continuous net reclassification improvement [NRI] [21]) for each of three aging measures were calculated, in comparison to those of the basic model with CA and gender only.

Since two of the three aging measures (PA.Accel and FI.Accel) outperformed relative to TL.Accel, we next tried to develop a new composite aging measure with better predictive utility by integrating aging measures at different hierarchical levels. Principal component analysis (PCA) was applied to PA.Accel and FI.Accel, and the first component (PC1) was defined as a new composite aging measure. We then performed the same analyses (i.e., K-M curves and Cox proportional hazards regression) to assess the predictive utility for all-cause mortality of PC1.

We applied linear regression to examine the responses to the lifestyle factors (i.e., smoking status, BMI, alcohol consumption, binge drinking status, PAQ, and HEI-2010) of PA.Accel, FI.Accel and PC1, the three showing significant predictive utilities of mortality in the previous analysis. Because PC1 was scaled, PA.Accel and FI.Accel were also scaled for comparability. We adjusted for CA and gender, and documented regression coefficients and 95% CI in these associations.

All analyses were conducted via R (version 4.0.3, 2020-10-10) and a two-sided p < 0.05 was considered to be statistically significant.

## Results

### Basic characteristics of study participants

The basic characteristics of 3249 participants are shown in **Table 1**. The mean CA of 3249 participants was 48.4 ± 17.8 years and around a third of them were old adults (≥ 60 years). Around half of the participants were females (50.8%). The proportions of non-Hispanic white, non-Hispanic black and Hispanic were 50.9%, 15.7%, and 30.6%, respectively. More than half of the participants didn’t go to college and only 18.2% received a bachelor’s degree or over. Half of the participants were never smokers, and 22.4% were current smokers. The proportions of participants at different alcohol consumption levels were similar. Around 13% of the participants reported being binge drinkers. Only 1.2% of participants were underweight and around 31% had normal weight. More than half reported performing physical activity less than one time per week. The mean HEI-2010 of the three tertiles group were 31.5, 45.7, and 62.3, respectively.

**Table 1.**
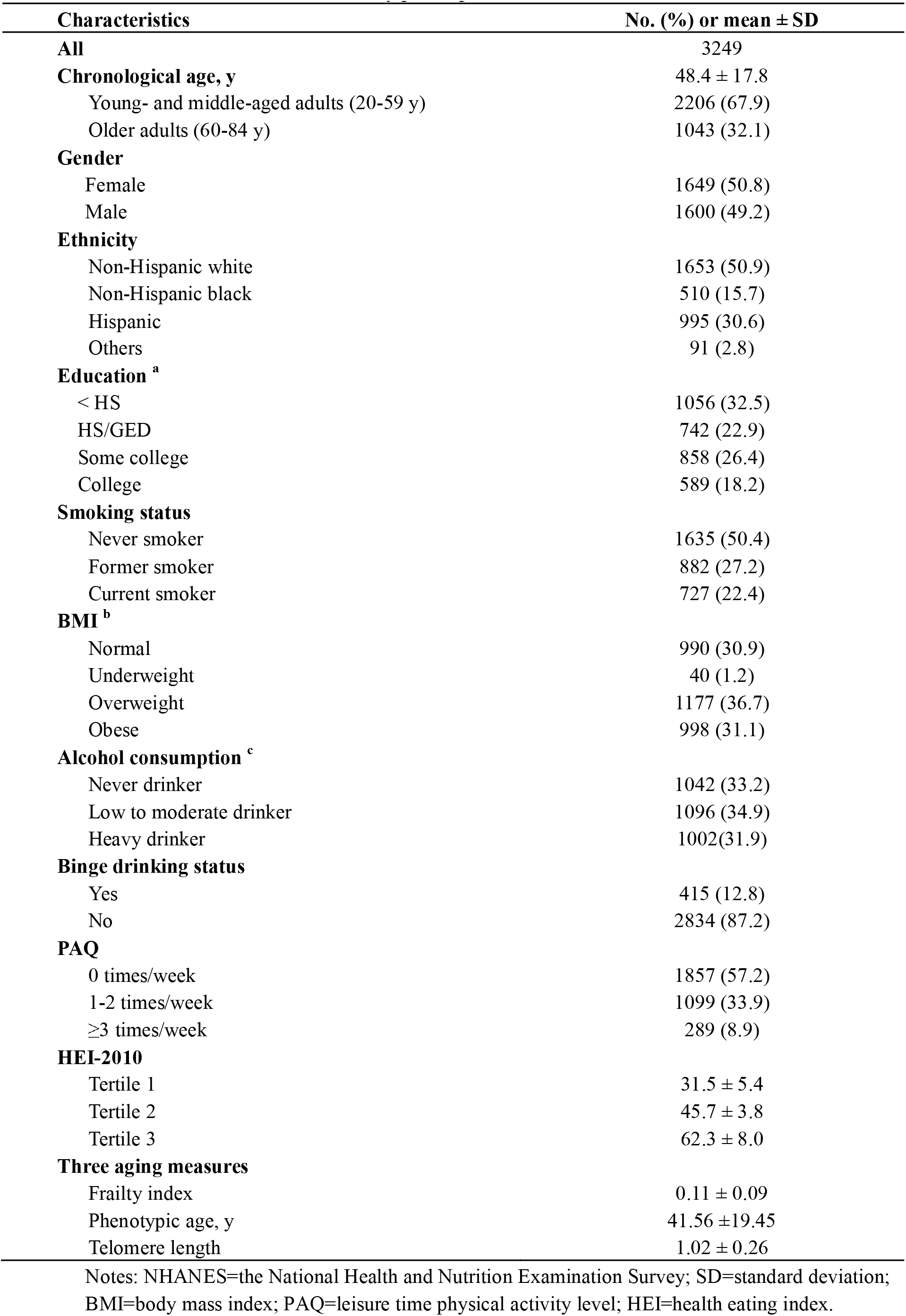
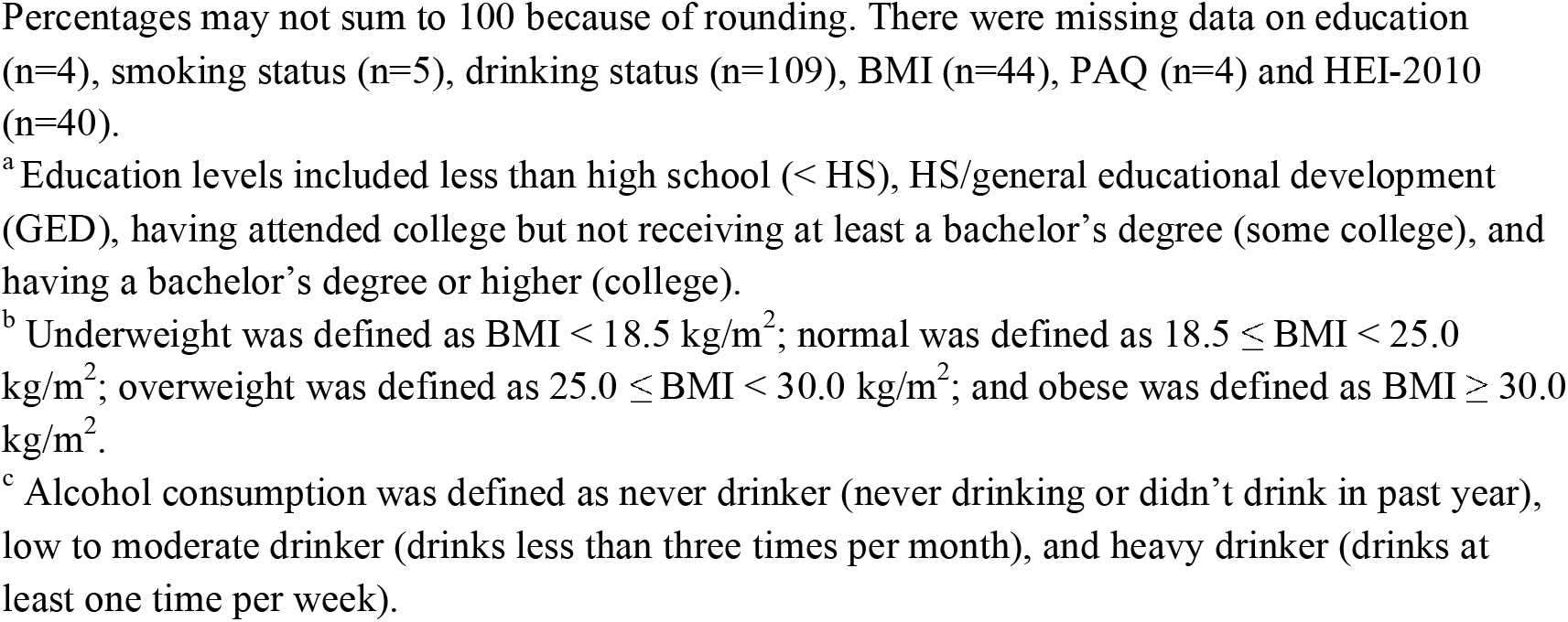
Characteristics of the study participants, NHANES 1999-2002.

### Were three aging measures correlated to CA?

As shown in **Fig. 1A**, all three aging measures significantly were correlated to CA. Among them, shorter TL was correlated to older CA with a Pearson correlation coefficient of -0.40, while the other two aging measures were positively correlated to CA. **Fig. 1B** illustrates the correlations after eliminating the effects of CA on aging measures by linear regression.

**Fig. 1.**
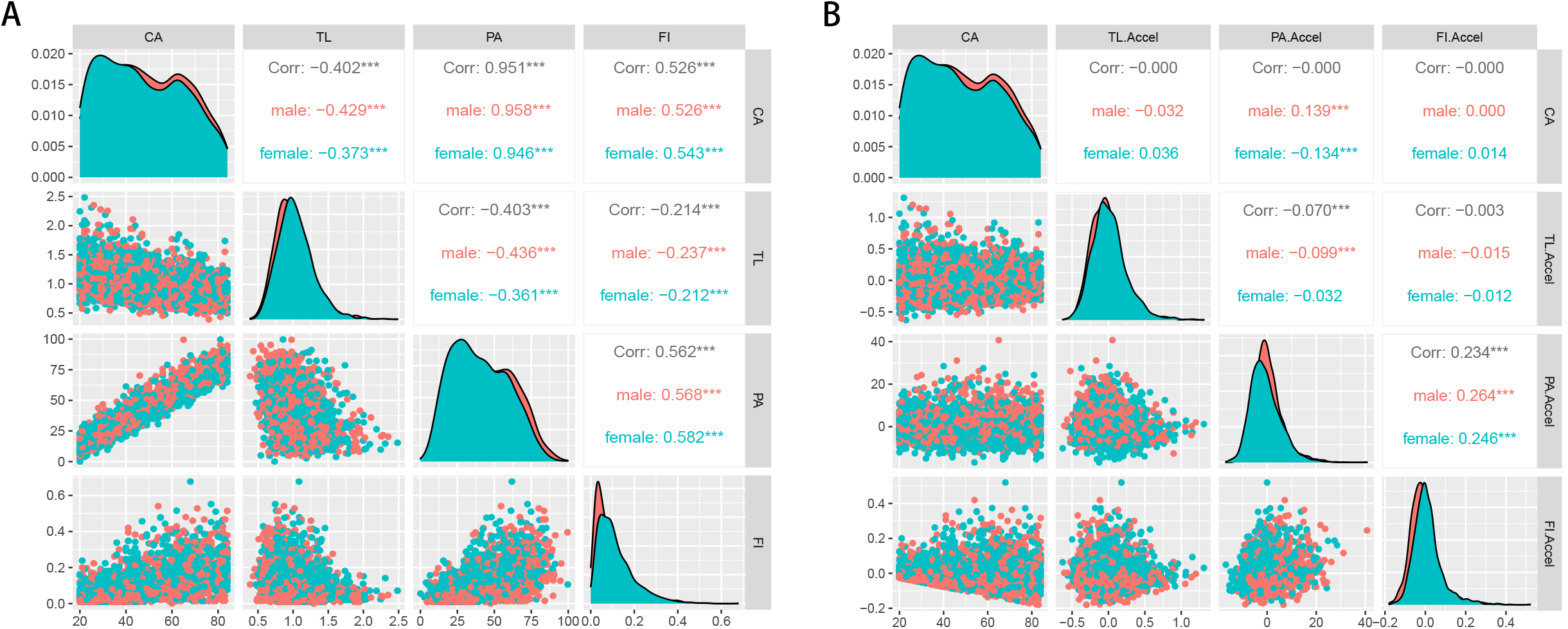
Correlations between three aging measures and chronological age. Note: CA=chronological age; TL=telomere length; PA=Phenotypic age; FI=frailty index; TL.Accel, PA.Accel and FI.Accel represent residuals from linear models when regressing TL, PA and FI on CA, respectively. ^***^ p < 0.001.

### Did three aging measures predict all-cause mortality?

**Fig. 2** presents the associations of the three aging measures with mortality. We found that PA.Accel (log-rank p < 0.001) and FI.Accel (log-rank p < 0.001), but not TL.Accel (log-rank p = 0.868), could identify participants at different risks of death. The similar results implied by Cox regression are shown in **Table 2**. According to the crude model (model 1), compared to phenotypically younger participants (PA.Accel < 0), phenotypically older participants (PA.Accel ≥ 0) had a 79% increase in mortality risk (HR = 1.79, 95% CI = 1.54-2.09). Similarly, compared to robust participants (FI.Accel ≥ 0), frail ones (FI.Accel < 0) had a 52% increase in mortality risk (HR = 1.52, 95% CI = 1.31-1.77). However, TL.Accel was found not to be significantly associated with mortality risk based on Cox regression (p = 0.868). After adjusting for covariates, these associations did not change substantially (models 2 and 3).

**Fig. 2.**
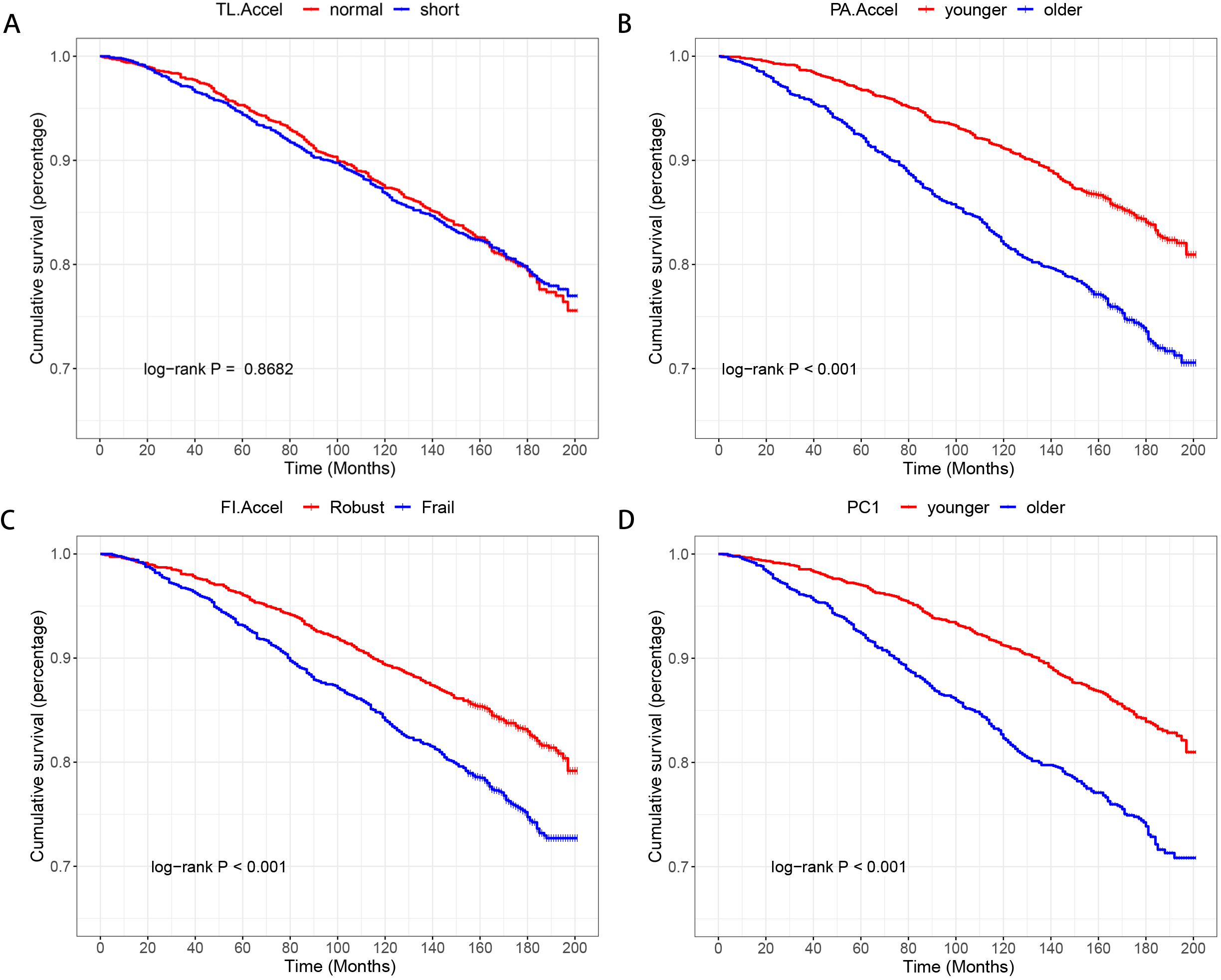
K-M curves of different aging measures for predicting all-cause mortality. Note: TL.Accel, PA.Accel and FI.Accel represent residuals from linear models when regressing telomere length, Phenotypic age and frailty index on chronological age, respectively. PC1 is the first component of PA.Accel and FI.Accel through the principle component analysis.

**Table 2.**
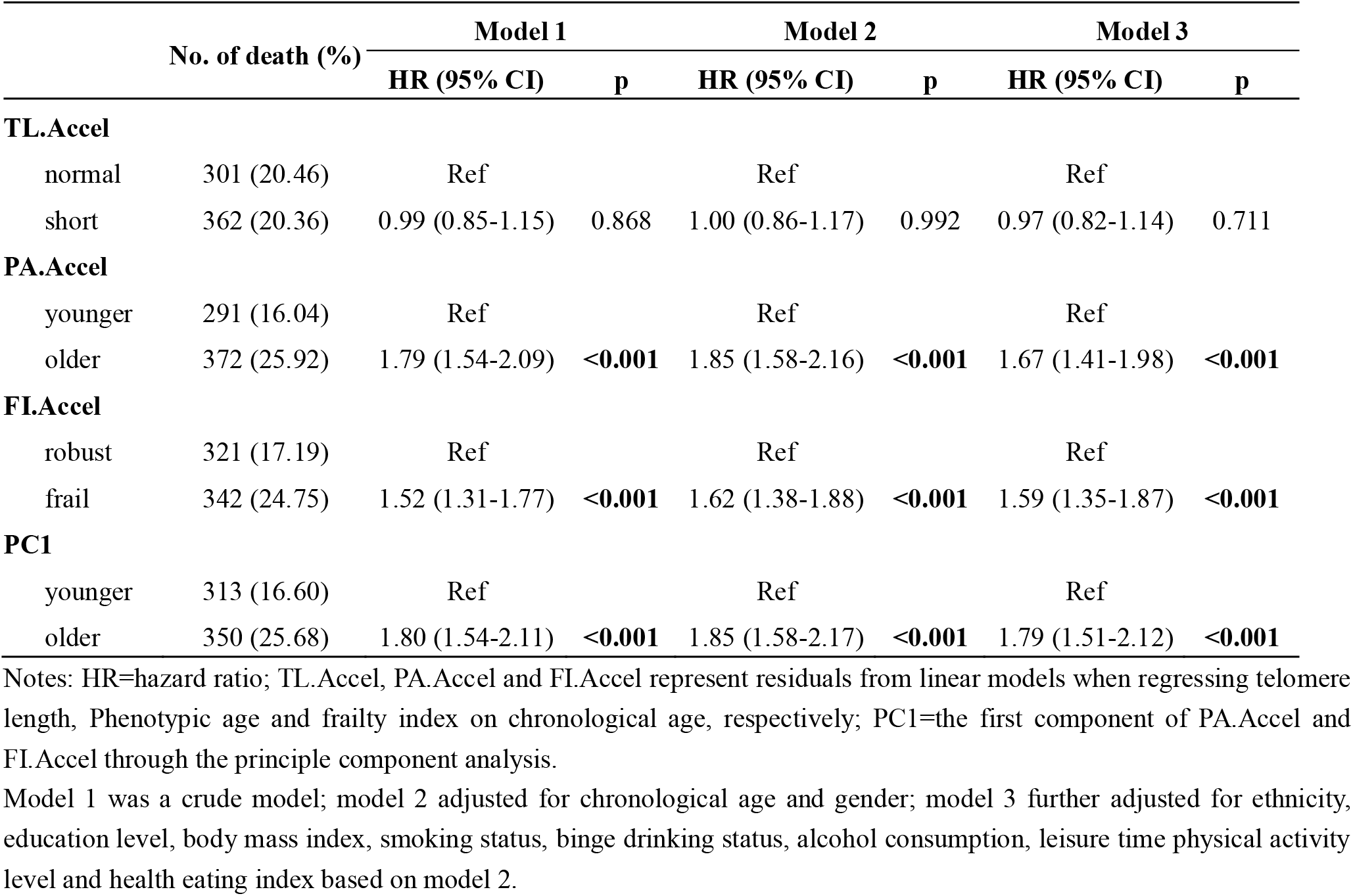
Associations of three aging measures with mortality.

### Did aging measures show additional predictive utilities than CA and gender?

**Fig. 3** exhibits the ROC curves for predicting mortality by different aging measures. Compared to the basic model (only CA and gender were included, AUC = 0.816), the model with PA.Accel or FI.Accel had higher predictive utility, evidenced by significantly increased AUC (PA.Accel: 0.829, p < 0.001; FI.Accel: 0.820, p = 0.067 in model 2), IDI (PA.Accel: 0.019, p < 0.001; FI.Accel: 0.010, p < 0.001 in model 2), and continuous NRI (PA.Accel: 0.193, p < 0.001; FI.Accel: 0.105, p < 0.001 in model 2). We did not observe that TL.Accel added significantly predictive utility. When adjusting for more covariates in the models (i.e., model 3), we observed similar patterns.

**Fig. 3.**
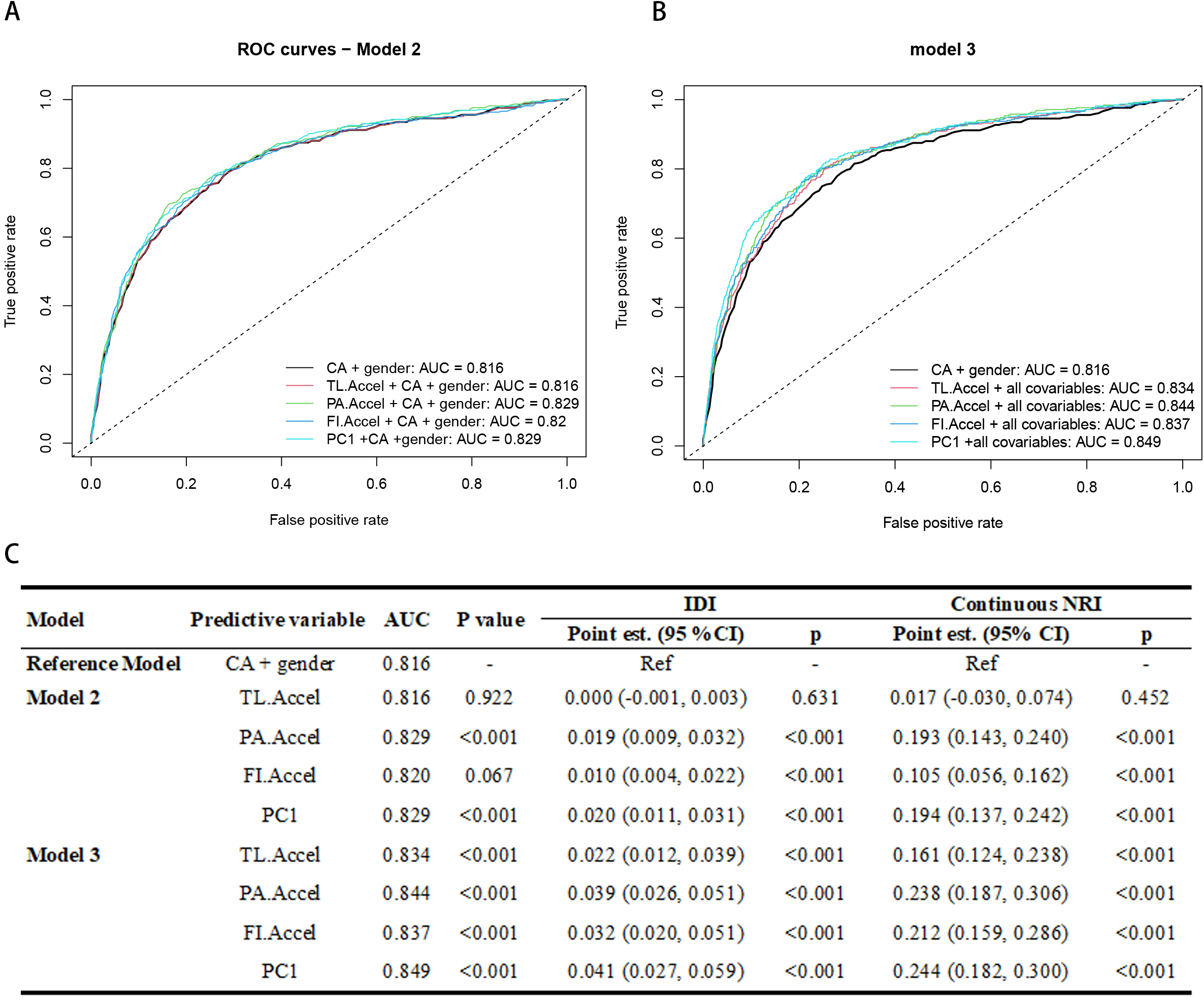
The predictive performance of different aging measures. Notes: CA=chronological age; TL=telomere length; PA=phenotypic age; FI=Frailty index; TL.Accel, PA.Accel and FI.Accel represent residuals from linear models when regressing telomere length, Phenotypic age and frailty index on CA, respectively; PC1=the first component of PA.Accel and FI.Accel through the principle component analysis; AUC=area under the curve; IDI=integrated discrimination improvement; NRI=net reclassification improvement; est.=estimation. Model 2 adjusted for CA and gender; Model 3 further adjusted for ethnicity, body mass index, education level, smoking status, alcohol consumption, binge drinking status, leisure time physical activity level and health eating index based on Model 2

### Can we develop a new composite aging measure?

Due to the inherent difference shared by aging measures at different hierarchical levels and the better predictive utility of PA.Accel and FI.Accel (relative to TL.Accel), we asked that whether we could develop a new composite aging measure with a better predictive utility by integrating aging measures at different hierarchical levels. Thus, PCA was applied to PA.Accel and FI.Accel and the scatter plot of PCA is shown in **Fig. S2** (**Supplementary file**). We found that PC1 accounted for 61.70% of the total variance and can be calculated as follows:

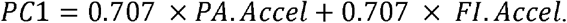

We then calculated PC1 for each participant. As shown in **Fig. 2D**, PC1 could identify participants at different risks of death (log-rank p < 0.001). Moreover, we found that PC1 outperformed each single aging measure (**Fig. 3C**) with larger AUC (0.829, p < 0.001, model 2), and greater increases of IDI (0.020, p < 0.001, model 2) and NRI (0.194, p < 0.001, model 2) compared to the basic model, and the pattern was more obvious in model 3.

### Are these aging measures responsive to modifiable lifestyle factors

**Fig. 4** presents results from linear regression to examine the association of modifiable lifestyle factors (i.e., smoking status, BMI, binge drinking status, alcohol consumption, PAQ, and HEI-2010) of PA.Accel, FI.Accel and PC1, the three showing significant predictive utilities of mortality in the previous analysis. Overall, PA.Accel, FI.Accel and PC1 were responsive to smoking status, BMI, alcohol consumption, and PAQ. For instance, compared to never smokers, current smokers had a significantly higher level of PA.Accel (β = 0.359, p <0.001) and FI.Accel (β = 0.231, p < 0.001). Interestingly, relative to PA.Accel and FI.Accel, PC1 showed stronger responses to almost all modifiable lifestyle factors (except for HEI-2010), with the largest absolute values of regression coefficients in these associations (**Table S2** in **Supplementary file**), indicating that the new composite aging measure might be more sensitive to modifiable lifestyle factors.

**Fig. 4.**
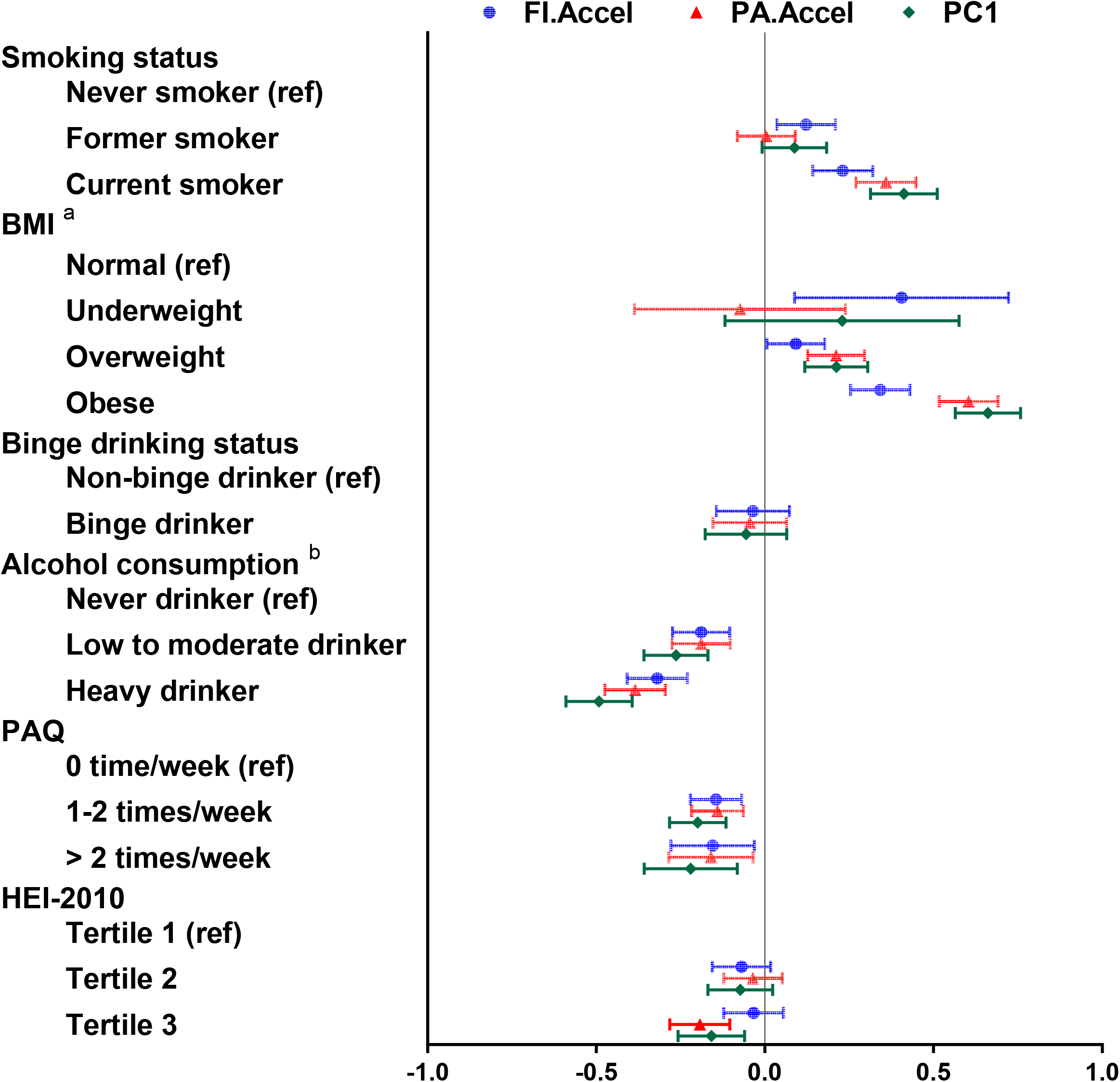
The responses of different aging measures to modifiable lifestyle factors. Notes: Coefficients (β) and 95% confidence intervals (CI) were calculated via linear regression adjusted for chronological age and gender. PA.Accel and FI.Accel represent residuals residuals from linear models when regressing telomere length, Phenotypic age and frailty index on chronological age, respectively. PC1=the first component of PA.Accel and FI.Accel through the principle component analysis; BMI=body mass index; PAQ=leisure time physical activity level; HEI= health eating index. ^a^ Alcohol consumption was defined as never drinker (never drinking or didn’t drink in past year), low to moderate drinker (drinks less than three times per month), and heavy drinker (drinks at least one time per week). ^b^ Underweight was defined as BMI < 18.5 kg/m^2^; normal was defined as 18.5 ≤ BMI < 25.0 kg/m^2^; overweight was defined as 25.0 ≤ BMI < 30.0 kg/m^2^; and obese was defined as BMI ≥ 30.0 kg/m^2^.

## Discussion

Based on the unique data from US NHANES, this study demonstrated that both PA and FI, but not TL, was significantly predictive of all-cause mortality. Building on the better performance of PA and FI, we integrated them to develop a new composite aging measure, which has been demonstrated to be predictive of mortality risk as well, even better than each single aging measure. Finally, we demonstrated that PA and FI, as well as the new composite aging measure, were responsive to some modifiable lifestyle factors, including smoking status, alcohol consumption, and PAQ. The findings, for the first time, provide a full picture of the predictive utility of mortality risk by three aging measures at three hierarchical levels and the response to modifiable lifestyle factors, with important implications for geroprotective programs.

The findings of the positive associations of PA and FI with all-cause mortality risk are consistent with previous studies [4, 22-25]. To date, the association of TL and mortality remains less conclusive in epidemiological studies [25-28], and the discrepancy may be partly explained by the differences among the study populations, and methods to measure TL [29]. Two studies based on the Dunedin birth cohort [25] and the National Health and Nutrition Examination Survey (NHANES) [27], respectively, considered TL and PA, and reported that TL was not consistently associated with multiple health span-related characteristics as compared to PA. However, FI was not considered in these two studies. The current study fills this knowledge gap by simultaneously evaluating the predictive utility of mortality risk by three aging measures at three hierarchical levels. The differences observed confirm that these aging measures did not necessarily reflect the same aging processes, as originally proposed by Ferrucci et al [1].

The increased predictive utility by PC1 relative to each single aging measure further demonstrated the differences shared by PA and FI. A similar finding was reported in a Canadian study in which FI-combined (the sum of the deficits in blood biomarkers and functional items) shows greater addition in the predictive utility of mortality relative to each single FI measure based on either blood biomarkers or functional terms [30]. The findings suggest that aging measures at phenotypic and functional levels might be complementary [8]. This indicates that integrating information across hierarchical levels may have the potential to develop better aging measures.

In addition to helping identify persons at risk, aging measures also serve as a potential endpoint for geroprotective programs. That being said, ideal aging measures should be responsive to risk factors [10]. In this study, PA and FI were found to meet this criterion since they were responsive to some modifiable lifestyle factors such as smoking status, BMI, alcohol consumption, and PAQ, which are largely consistent with previous studies [31-33]. More interestingly, the new composite aging measure we developed, PC1, was strongly responsive to the same set of modifiable lifestyle factors, highlighting its qualification as an aging measure.

Our findings have important implications. First, the predictive utility of mortality risk by these aging measures (PA, FI, and PC1) suggests that we could identify vulnerable persons at risk of early death. Together with the fact that they were responsive to modifiable lifestyle factors, it seems that lifestyle-targeted interventions may have the potential to slow aging and further reduce the burden of early death. Finally, it is promising to use these aging measures (particularly PC1) to test the effectiveness of geroprotective programs in human beings. Application of aging measures is more practical and feasible in comparison to previous approaches using endpoints such as death, and the occurrence of chronic diseases, the latter requiring a long time of follow-up and high expenditures.

The present study has several strengths. First, we compared aging measures at three hierarchical levels in the same population, which is scarce in the literature. Second, the three aging measures we adopted in this study are widely recognized in the literature. We also acknowledge limitations in this study. First, the findings were based on the US population and thus may not be generalizable to other populations from different countries. Second, due to the unavailability of repeated measurements of these aging measures, we were unable to evaluate the associations between the rate of changes in aging measures and mortality risk. Third, only one aging measure at each hierarchical level was considered, in particular, only TL at the biological level. In recent years, DNA methylation age has been widely demonstrated as a promising aging measure [34-38]; however, it was not available in the NHANES data. In moving forward, with more aging measures available, a more comprehensive picture of aging would be forthcoming.

## Conclusions

Our study demonstrates that both phenotypic (i.e., PA) and functional (i.e., FI) aging measures can capture mortality risk and respond to modifiable lifestyle factors, despite their inherent differences. Furthermore, the PC1 that integrated phenotypic and functional aging measures outperforms in predicting mortality risk in comparison with each single aging measure, and strongly responds to modifiable lifestyle factors. The findings suggest the complementary of aging measures at different hierarchical levels and underscore the need to involve multi-level information when quantifying aging. The findings also highlight the potential of lifestyle-targeted interventions as geroprotective programs.

## Data Availability

All data produced are available online at: https://www.cdc.gov/nchs/nhanes/index.htm

## Contributions

Conceptualization was proposed by Jingyun Zhang, Xifeng Wu, and Zuyun Liu. The methodological design was conducted by Jingyun Zhang and Zuyun Liu. Data collection and preparation were performed by Chen Chen, Xingqi Cao, and Zuyun Liu. Data analyses were conducted by Jingyun Zhang and Zuyun Liu. The results were interpreted by Junhua Xiao and Liyuan Han. The first draft of the manuscript was written by Jingyun Zhang, Xingqi Cao, Liu He, and Ziyang Ren. All authors reviewed and edited the manuscript. Xifeng Wu and Zunyun Liu provided overall supervision.

## Acknowledgment

We thank all participants who attended the National Health and Nutrition Examination Survey (NHANES).

## Data availability

NHANES data are publicly available (https://www.cdc.gov/nchs/nhanes/index.htm).

## Ethics declarations

### Ethics approval

NHANES is approved by the National Center for Health Statistics Research Ethics Review Board.

### Consent to participate

All participants provided informed consent.

## References

1. Ferrucci, L., et al., Time and the Metrics of Aging. Circ Res, 2018. 123(7): p. 740–744.

2. Horvath, S. and K. Raj, DNA methylation-based biomarkers and the epigenetic clock theory of ageing. Nat Rev Genet, 2018. 19(6): p. 371–384.

3. Bell, C.G., et al., DNA methylation aging clocks: challenges and recommendations. Genome Biol, 2019. 20(1): p. 249.

4. Liu, Z., et al., A new aging measure captures morbidity and mortality risk across diverse subpopulations from NHANES IV: A cohort study. PLoS Med, 2018. 15(12): p. e1002718.

5. Mitnitski, A.B., A.J. Mogilner, and K. Rockwood, Accumulation of deficits as a proxy measure of aging. ScientificWorldJournal, 2001. 1: p. 323–36.

6. Rockwood, K. and A. Mitnitski, Frailty in relation to the accumulation of deficits. J Gerontol A Biol Sci Med Sci, 2007. 62(7): p. 722–7.

7. Rockwood, K., et al., A global clinical measure of fitness and frailty in elderly people. CMAJ, 2005. 173(5): p. 489–95.

8. Li, X., et al., Longitudinal trajectories, correlations and mortality associations of nine biological ages across 20-years follow-up. Elife, 2020. 9.

9. Lehallier, B., et al., Undulating changes in human plasma proteome profiles across the lifespan. Nat Med, 2019. 25(12): p. 1843–1850.

10. Justice, J.N. and S.B. Kritchevsky, Putting epigenetic biomarkers to the test for clinical trials. Elife, 2020. 9.

11. Simpson, R.J., et al., Exercise and the aging immune system. Ageing Res Rev, 2012. 11(3): p. 404–20.

12. Brown, J.D., et al., Association between a Deficit Accumulation Frailty Index and Mobility Outcomes in Older Adults: Secondary Analysis of the Lifestyle Interventions and Independence for Elders (LIFE) Study. J Clin Med, 2020. 9(11).

13. Huang, C.H., et al., Effect of various exercises on frailty among older adults with subjective cognitive concerns: a randomised controlled trial. Age Ageing, 2020. 49(6): p. 1011–1019.

14. Cawthon, R.M., Telomere measurement by quantitative PCR. Nucleic Acids Res, 2002. 30(10): p. e47.

15. Needham, B.L., et al., Socioeconomic status, health behavior, and leukocyte telomere length in the National Health and Nutrition Examination Survey, 1999-2002. Soc Sci Med, 2013. 85: p. 1–8.

16. Levine, M.E., et al., An epigenetic biomarker of aging for lifespan and healthspan. Aging (Albany NY), 2018. 10(4): p. 573–591.

17. Blodgett, J.M., et al., A frailty index from common clinical and laboratory tests predicts increased risk of death across the life course. Geroscience, 2017. 39(4): p. 447–455.

18. Statistics, N.C.f.H., Office of Analysis and Epidemiology, Public-use Linked Mortality File, 2015. Hyattsville, Maryland. (Available at the following address: https://www.cdc.gov/nchs/data-linkage/mortality-public.htm).

19. Guenther, P.M., et al., The Healthy Eating Index-2010 is a valid and reliable measure of diet quality according to the 2010 Dietary Guidelines for Americans. J Nutr, 2014. 144(3): p. 399–407.

20. Kamarudin, A.N., T. Cox, and R. Kolamunnage-Dona, Time-dependent ROC curve analysis in medical research: current methods and applications. BMC Med Res Methodol, 2017. 17(1): p. 53.

21. Pencina, M.J., et al., Evaluating the added predictive ability of a new marker: from area under the ROC curve to reclassification and beyond. Stat Med, 2008. 27(2): p. 157–72; discussion 207-12.

22. Kojima, G., S. Iliffe, and K. Walters, Frailty index as a predictor of mortality: a systematic review and meta-analysis. Age Ageing, 2018. 47(2): p. 193–200.

23. Fan, J., et al., Frailty index and all-cause and cause-specific mortality in Chinese adults: a prospective cohort study. Lancet Public Health, 2020. 5(12): p. e650–e660.

24. Liu, Z., et al., Associations of genetics, behaviors, and life course circumstances with a novel aging and healthspan measure: Evidence from the Health and Retirement Study. PLoS Med, 2019. 16(6): p. e1002827.

25. Belsky, D.W., et al., Eleven Telomere, Epigenetic Clock, and Biomarker-Composite Quantifications of Biological Aging: Do They Measure the Same Thing? Am J Epidemiol, 2018. 187(6):p, 12201230.

26. Wang, Q., et al., Telomere Length and All-Cause Mortality: A Meta-analysis. Ageing Res Rev, 2018. 48: p. 11–20.

27. Hastings, W.J., I. Shalev, and D.W. Belsky, Comparability of biological aging measures in the National Health and Nutrition Examination Study, 1999-2002. Psychoneuroendocrinology, 2019. 106: p. 171–178.

28. Cawthon, R.M., et al., Association between telomere length in blood and mortality in people aged 60 years or older. Lancet, 2003. 361(9355): p. 393–5.

29. Sanders, J.L. and A.B. Newman, Telomere length in epidemiology: a biomarker of aging, age-related disease, both, or neither? Epidemiol Rev, 2013. 35(1): p. 112–31.

30. Howlett, S.E., et al., Standard laboratory tests to identify older adults at increased risk of death. BMC Med, 2014. 12: p. 171.

31. Kresovich, J.K., et al., Associations of Body Composition and Physical Activity Level With Multiple Measures of Epigenetic Age Acceleration. Am J Epidemiol, 2021. 190(6): p. 984–993.

32. Peng, H., et al., Combined healthy lifestyle score and risk of epigenetic aging: a discordant monozygotic twin study. Aging (Albany NY), 2021. 13(10): p. 14039–14052.

33. Ng, T.P., et al., Socio-Environmental, Lifestyle, Behavioural, and Psychological Determinants of Biological Ageing: The Singapore Longitudinal Ageing Study. Gerontology, 2020. 66(6): p. 603–613.

34. Chen, B.H., et al., DNA methylation-based measures of biological age: meta-analysis predicting time to death. Aging (Albany NY), 2016. 8(9): p. 1844–1865.

35. Marioni, R.E., et al., DNA methylation age of blood predicts all-cause mortality in later life. Genome Biol, 2015. 16(1): p. 25.

36. Levine, M.E., et al., Epigenetic age of the pre-frontal cortex is associated with neuritic plaques, amyloid load, and Alzheimer’s disease related cognitive functioning. Aging (Albany NY), 2015. 7(12): p. 1198–211.

37. Martin-Herranz, D.E., et al., Screening for genes that accelerate the epigenetic aging clock in humans reveals a role for the H3K36 methyltransferase NSD1. Genome Biol, 2019. 20(1): p. 146.

38. Zheng, Y., et al., Blood Epigenetic Age may Predict Cancer Incidence and Mortality. EBioMedicine, 2016. 5: p. 68–73.

