## Supplementary material for "Predictive utility of mortality by aging measures at different hierarchical levels and the response to modifiable lifestyle factors: Implications for geroprotective programs": Table S1 (Supplementary file)

**Table S1. Thirty-six items to construct frailty index in NHANES [1]**

| **Item No.** | **deficit** |
| --- | --- |
| 1 | General hearing |
| 2 | Has high blood pressure |
| 3 | Self-reported poor health |
| 4 | Has diabetes |
| 5 | Frequency of healthcare use |
| 6 | Health compared to 1 year ago |
| 7 | Overnight hospital stays |
| 8 | Leaked/lost control of urine |
| 9 | Weak/failing kidneys |
| 10 | Has angina/angina pectoris |
| 11 | Has cancer |
| 12 | Has arthritis |
| 13 | Has heart attack |
| 14 | Has heart disease |
| 15 | Has thyroid condition |
| 16 | Has stroke |
| 17 | Has broken hip |
| 18 | Has osteoporosis |
| 19 | Confusion or inability to remember things |
| 20 | Difficulty in attending social events |
| 21 | Difficulty in dressing yourself |
| 22 | Difficulty in getting in and out of bed |
| 23 | Difficulty in grasping/holding small objects |
| 24 | Difficulty in lifting or carrying |
| 25 | Difficulty in managing money |
| 26 | Difficulty in preparing meals |
| 27 | Difficulty in pushing or pulling large objects |
| 28 | Difficulty in standing up from the armless chair |
| 29 | Difficulty in stooping, crouching, kneeling |
| 30 | Difficulty in using fork and knife |
| 31 | Difficulty in walking between rooms on the same floor |
| 32 | Have you taken or used any prescription medicines in the past month? |
| 33 | Cough regularly |
| 34 | General vision |
| 35 | Cataract operation |
| 36 | Difficulty in seeing steps/curbs in dim light |

Notes: NHANES=the National Health and Nutrition Examination Survey.

**Reference:**

[1] Blodgett, J.M., et al., *A frailty index from common clinical and laboratory tests predicts increased risk of death across the life course.* Geroscience, 2017. **39**(4): p. 447-455.

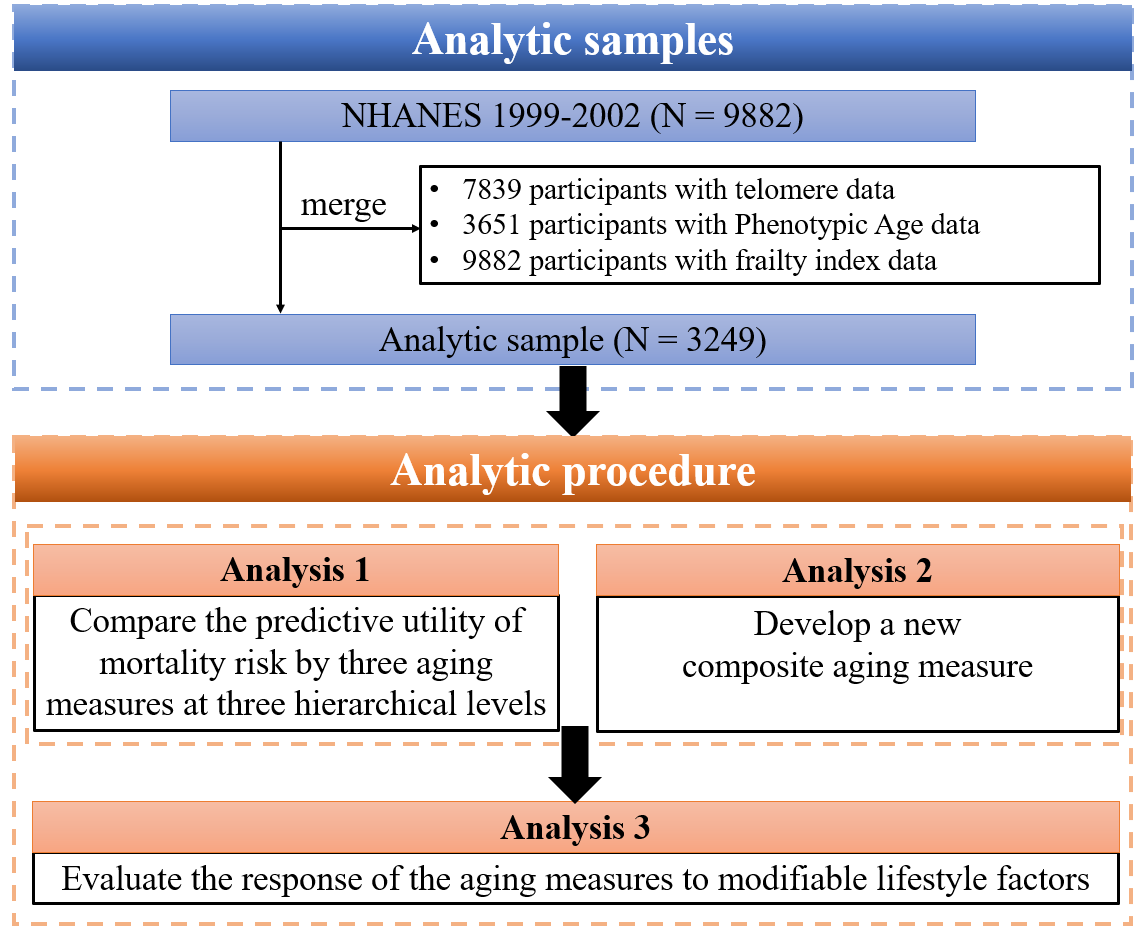

**Fig. S1. Analytic roadmap of this study**

Notes: NHANES=the National Health and Nutrition Examination Survey.

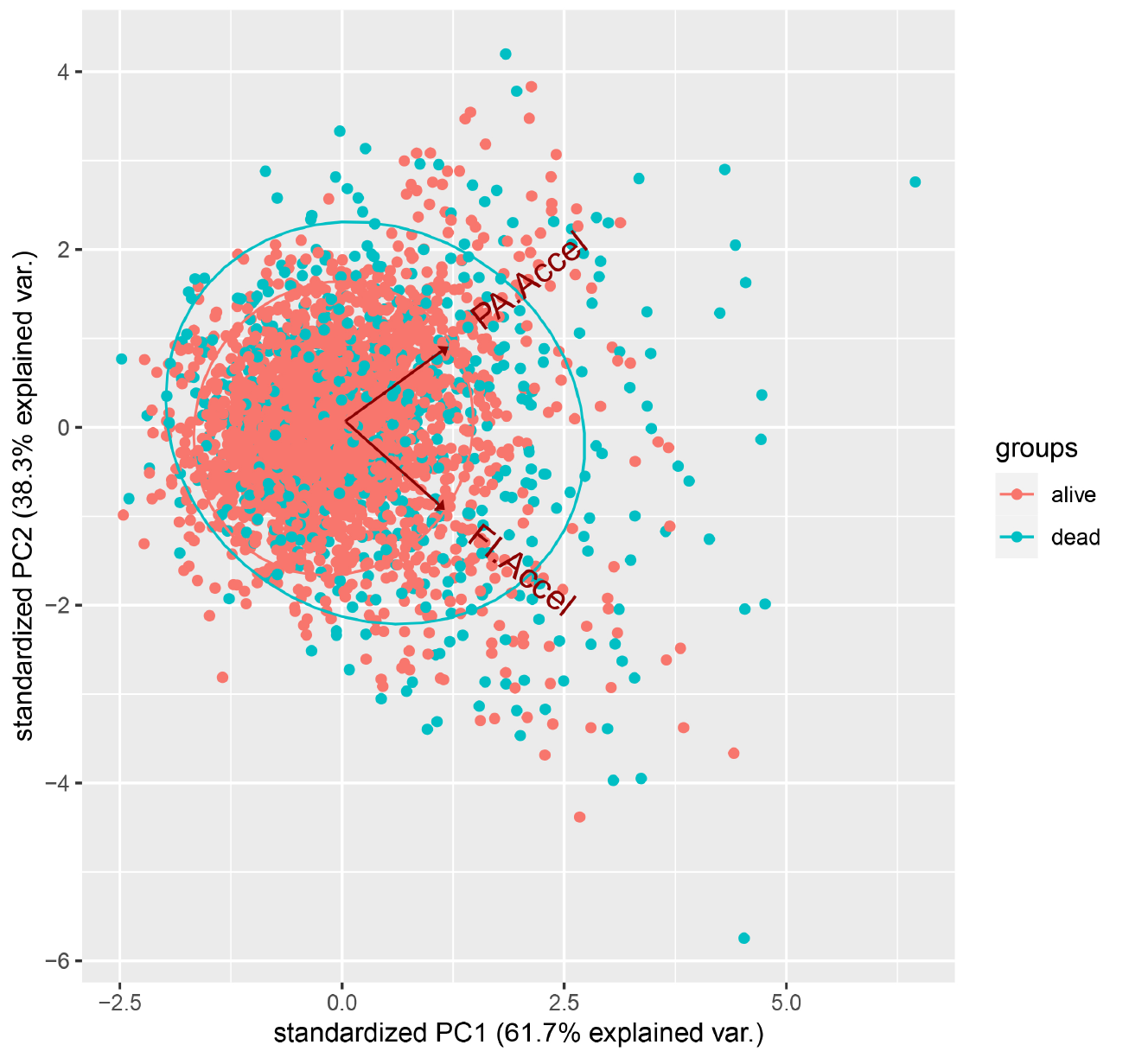

**Fig. S2. Scatter plot of principal component analysis of accelerations of Phenotypic Age and Frailty Index**

Notes: PA.Accel=residual from linear regression when regressing Phenotypic age on chronological age; FI.Accel=residual from linear regression when regressing frailty index on chronological age; PC1=the first component of PA.Accel and FI.Accel through the principal component analysis; PC2=the second component of PA.Accel and FI.Accel through the principal component analysis.

**Table S2. Responses of different aging measures to modifiable lifestyle factors**

| **Risk factors** | | **PA.Accel** | | |  | **FI.Accel** | | |  | **PC1** | | |
| --- | --- | --- | --- | --- | --- | --- | --- | --- | --- | --- | --- | --- |
|  |  | **β ^a^** | **95% CI** | **P-value** |  | **β** | **95% CI** | **P-value** |  | **β** | **95% CI** | **P-value** |
| **Smoking status** | | | | | | | | | | | | |
|  | Never smoker | ref | | | | | | | | | | |
|  | Former smoker | 0.004 | (-0.082, 0.090) | 0.928 |  | 0.122 | (0.036, 0.209) | **0.006** |  | 0.088 | (-0.008, 0.183) | 0.073 |
|  | Current smoker | 0.359 | (0.270, 0.448) | **<0.001** |  | 0.231 | (0.141, 0.320) | **<0.001** |  | 0.412 | (0.313, 0.511) | **<0.001** |
| **BMI categories ^b^** | | | | | | | | | | | | |
|  | Normal | ref | | | | | | | | | | |
|  | Underweight | -0.074 | (-0.387, 0.239) | 0.643 |  | 0.405 | (0.088, 0.722) | **0.012** |  | 0.229 | (-0.118, 0.576) | 0.196 |
|  | Overweight | 0.211 | (0.127, 0.295) | **<0.001** |  | 0.092 | (0.006, 0.177) | **0.035** |  | 0.212 | (0.118, 0.305) | **<0.001** |
|  | Obese | 0.604 | (0.517, 0.691) | **<0.001** |  | 0.342 | (0.254, 0.430) | **<0.001** |  | 0.661 | (0.565, 0.758) | **<0.001** |
| **Alcohol consumption ^c^** | | | | | | | | | | | | |
|  | Never drinker | ref | | | | | | | | | | |
|  | Low to moderate drinker | -0.189 | (-0.275, -0.103) | **<0.001** |  | -0.189 | (-0.274, -0.104) | **<0.001** |  | -0.264 | (-0.358, -0.169) | **<0.001** |
|  | Heavy drinker | -0.385 | (-0.474, -0.295) | **<0.001** |  | -0.319 | (-0.408, -0.230) | **<0.001** |  | -0.492 | (-0.590, -0.393) | **<0.001** |
| **Binge drinking status** | | | | | | | | | | | | |
|  | No | ref | | | | | | | | | | |
|  | Yes | -0.045 | (-0.154, 0.065) | 0.423 |  | -0.035 | (-0.144, 0.073) | 0.523 |  | -0.056 | (-0.177, 0.065) | 0.365 |
| **PAQ** | | | | | | | | | | | | |
|  | 0 times/week | ref | | | | | | | | | | |
|  | 1-2 times/week | -0.140 | (-0.217, -0.064) | **<0.001** |  | -0.145 | (-0.221, -0.069) | **<0.001** |  | -0.199 | (-0.283, -0.115) | **<0.001** |
|  | ≥3times/week | -0.160 | (-0.285, -0.035) | **0.012** |  | -0.155 | (-0.279, -0.031) | **0.015** |  | -0.220 | (-0.358, -0.082) | **0.002** |
| **HEI-2010** | | | | | | | | | | | | |
|  | Tertile 1 | ref | | | | | | | | | | |
|  | Tertile 2 | -0.035 | (-0.122, 0.051) | 0.426 |  | -0.069 | (-0.156, 0.017) | 0.115 |  | -0.073 | (-0.169, 0.023) | 0.138 |
|  | Tertile 3 | -0.193 | (-0.282, -0.104) | **<0.001** |  | -0.034 | (-0.122, 0.055) | 0.456 |  | -0.159 | (-0.257, -0.060) | **0.002** |

Notes: BMI=body mass index; PAQ=leisure time physical activity level; HEI=health eating index; PA.Accel=residual from linear regression when regressing Phenotypic age on chronological age; FI.Accel=residual from linear regression when regressing frailty index on chronological age; PC1=the first component of PA.Accel and FI.Accel through the principal component analysis;

^a^ Coefficients (β) and 95% confidence intervals were calculated via linear regression after adjusted for chronological age and gender.

^b^ Underweight was defined as BMI < 18.5 kg/m^2^; normal was defined as 18.5 ≤ BMI < 25.0 kg/m^2^; overweight was defined as 25.0 ≤ BMI < 30.0 kg/m^2^; and obese was defined as BMI ≥ 30.0 kg/m^2^.

^c^ Alcohol consumption was defined as never drinker (never drinking or didn’t drink in the past year), low to moderate drinker (drinks less than three times per month), and heavy drinker (drinks at least one time per week).
